# Using Machine Learning to Examine Pre-Transplant Factors Influencing *De novo* HLA-Specific Antibody Development Post-Kidney Transplant

**DOI:** 10.1101/2024.10.28.24315920

**Authors:** Alex Rothwell, George Nita, Matthew Howse, Dan Ridgway, Abdul Hammad, Sanjay Mehra, Andrew R Jones, Petra Goldsmith

## Abstract

The development of *de novo* donor-specific antibodies (DSAs) against HLA is associated with premature graft failure in kidney transplantation. However, rates and factors influencing *de novo* DSA formation vary widely across the literature. We aimed to identify pre-transplant factors influencing the development of *de novo* HLA-specific antibodies following kidney transplantation using machine learning.

Data from 460 kidney transplant recipients at a single centre between 2009-2014 were analysed. Pre-transplant variables were collected, and post-transplant sera were screened for HLA antibodies. Positive samples were investigated using Single Antigen Bead (SAB) testing. Machine learning models (Classification and Regression Trees, Random Forest, XGBoost, CatBoost) were trained on a training set of pre-transplant data to predict *de novo* DSA formation, with and without SMOTE oversampling. Model performance was evaluated on an independent testing set using F1 scores, and feature importance was assessed using SHAP.

In the full cohort analysis, XGBoost models performed the best, with F1 scores of 0.54-0.59 without SMOTE and 0.72-0.79 with SMOTE. The strongest predictors were pre-transplant HLA antibodies, number of kidney transplants, cold ischemia time (CIT), recipient age and female gender. SHAP dependence plots showed that pre-existing HLA antibodies and past transplants increased the risk of *de novo* DSA development. In the unsensitised subgroup analysis, model performance was poor.

Machine learning models can be used to identify pre-transplant risk factors for *de novo* HLA-specific antibody development in kidney transplantation. Monitoring and risk-stratifying patients based on these factors may help guide preventive immunological strategies and recipient selection to improve long-term allograft outcomes.

**Translational statement:** This study identified pre-transplant risk factors for the development of *de novo* HLA-specific antibody in kidney transplantation. Monitoring and risk-stratifying patients based on these factors may help guide preventive immunological strategies and recipient selection to improve long-term allograft outcomes.

## Introduction

*De novo* donor specific antibody (DSA) directed against HLA is associated with premature graft failure in renal transplantation ^1–11^. The clinical picture can vary: grafts may experience acute dysfunction, gradual decline, and even demonstrate no effect despite persistent DSA.

The rates of any *de novo* HLA-specific antibody (DSA and non-DSA) post-transplant vary enormously, ranging from 1.6-60% ^12^. There is evidence that developing *de novo* DSA, but not non-donor *de novo* HLA-specific antibody, has poorer outcomes ^13^. However, Worthington et al identified that 50.9% of patients who lost their allografts produced HLA-specific antibody, compared to only 1.6% in those with a functioning allograft, and that 60% of those with graft loss and HLA-specific antibody developed antibody prior to graft loss ^3^, not all of them donor-specific. Thus, all *de novo* HLA-specific antibody appears to be implicated in adverse outcomes in renal transplantation.

Several studies have shown that 7-11% of renal transplant recipients had circulating HLA-specific antibody but only 4-5.5% of these were post-transplant DSA ^4,5^. The overall frequency of HLA-specific antibody development in unsensitised pre-transplant patients was described as 14.7% ^9^. Graft failure rates were higher in the *de novo* antibody group at 8.6% versus 3% in the group without antibodies^9^.

To understand which pre-transplant factors influence the development of *de novo* HLA-specific antibody following kidney transplant, we devised a study using a combination of univariate statistics and machine learning. Machine learning (ML) has been considered to be unhelpful in predicting post-transplant outcomes, partly due to their “black-box” nature ^14^. However, artificial intelligence (AI) and ML integration in enhancing perioperative care in transplantation, from donor and recipient selection to post-operative prevention of complications, and graft outcome prediction, remains a current research focus ^15^. An increasing number of studies show promising applications of ML in developing prediction models in kidney transplantation. Ravindhran *et al.* published a recent systematic literature review and meta-analysis examining the current application of ML in kidney transplantation concluding that hybrid ML models, variations of Support Vector Machines (SVM), Random Forest (RF)-based models appear to perform the best in prediction of graft survival ^16^. Principal component analysis (PCA)-derived algorithms can be used successfully to identify new patterns of reactivity that differ from patients’ historic HLA antibody pattern (Sn 100%, 95% CI, 73.54-100%, Sp 75%, 95% CI, 56.30%-92.54%) ^17^. Growing evidence supports utilising ML in decision-making in kidney transplantation. Recurrent Neural Network (RNN) models can provide support in determining the most optimal donor-recipient pairing and informed-decision making ^18^. Explainable AI (XAI) are systems that offer explanations for ML models’ outputs, aiming to improve interpretability, therefore increasing trust and confidence in predictions. This has been of particular interest in the medical field, where ML has the potential to benefit patients and health services, though with serious consequences if models perform poorly ^19^. Our study aims to add to the body of literature supporting the integration of ML, by using XAI to identify pre-transplant variables leading to *de novo* HLA-specific antibody development.

Therefore, we attempt to describe the emergence of *de novo* HLA-specific antibody following renal transplant in an uncensored renal transplant cohort and an unsensitised subset, to examine the pre-transplant factors implicated in its development.

## Materials and Methods

### Data collection

Ethical approval was granted from the Health Research Authority National Research Ethics Service, study number 11/NW/0279 and the local Research and Development Department, who acted as sponsors, reference number 4049. The study participants had received a kidney transplant at the Royal Liverpool University Hospital between 2009 and 2014. Following informed consent, all samples available for each patient at 2 weeks, 1 month, 3 months, 6 months, 9 months, 12 months and subsequently yearly following transplant underwent a Labscreen® Mixed screening assay (LSM12, Lot 17) to broadly determine the presence or absence of Class I or Class II HLA-specific antibodies. The assay was carried out as per manufacturers specification and with the same methodology we have previously described ^20,21^. Samples with a positive result, defined as mean fluorescence intensity (MFI) ≥500) for class I and/or class II, were retained for single antigen bead (SAB) testing. For each patient, only the first positive sample for class I and class II were tested. The technique for testing with SAB was the same as that used for the Labscreen® Mixed assay, however, the threshold for a positive result was locally set at a normalised MFI of ≥2000 and the beads were specific for single Class I: HLA Labscreen® Single Antigen HLA Class I – Combi (lot numbers 9 and 10) and Class II: LABScreen® Single Antigen HLA Class II Group 1 (Lots 10 and 11).

### Statistical Analyses

Clinical data, including recipient demographic data about all eligible participants, was collated from databases and software in the Royal Liverpool University Hospital and are listed in Table 1. Statistical analysis was performed in Microsoft Excel. For univariate analysis, categorical variables were analysed with Chi Square tests and non-parametric continuous variables were analysed with Mann Whitney U tests. P-values were adjusted using the Benjamini-Hochberg procedure to control for family-wise error. None of the data was considered to be normally distributed. Ethnicity was eliminated from multivariate and machine learning analysis due to the very small portion of Black, Asian and Minority Ethnic groups represented and intrinsic diversity in the subgroups precluding their fusion.

**Table 1:** Pre- Transplants Variables used for statistical and machine learning analyses.

|  |
| --- |
| Age |
| Gender |
| Ethnicity |
| Mode of Dialysis |
| Number of kidney transplants |
| HLA Mismatches (A, B, Cw, DR, DQ) |
| Type of transplant |
| Amino Acid Mismatch |
| Cold Ischaemic Time (CIT) |
| Induction Immunosuppression |
| Pre-Transplant HLA-Specific<br>Antibody |

Two iterations of machine learning analysis were performed a) using the whole cohort b) using the cohort of unsensitised pre-transplant patients. The reason for performing two iterations is due to significant biases in both analyses i.e. in the total cohort, the results were heavily influenced by pre-formed HLA-specific antibody which appeared to account for almost all of the effect of the analysis, whereas in the unsensitised subset, analysis was limited in credibility by the small number of *de novo* HLA antibodies formed post-transplant. Thus, examining the same analysis from the two different cohorts aimed to provide evidence that addressed at least some of the foreseen bias encountered.

### Machine Learning

Four different classification algorithms were trained on the pre-transplant factors listed to predict whether transplant patients would or would not develop *de novo* HLA-specific antibody. Classification and Regression Trees (CART), Random Forest (RF), CatBoost (CB) and XGBoost (XGB) are tree-based algorithms that can be used for classification. CART are binary trees that mathematically determine the best split on single features. Using the classifiers for prediction involves following the series of rules generated by the tree, e.g. if number of kidney transplants is > 0 and gender is female, then “does not develop *de novo* HLA-specific antibody post-transplant” may be predicted. RFs create an ensemble of trees, each tree is varied due to having been limited to searching over a random subset of features on a random sample of training data when generating decision rules, with the output being the class voted by most trees ^22^. XGB is a gradient boosting algorithm that works by combining a series of weak tree classifiers which sequentially aim to further minimise the training error of the previous tree, ultimately resulting in a strong classifier ^23^. CB is also a gradient boosting algorithm, similar to XGB. Though it has been shown to outperform XGB at times, especially in the presence of categorical features, which CB is capable of handling natively without requiring them to be encoded numerically. For example, through the use of one hot encoding which involves creating a new column for each categorical value then assigning a binary value to represent presence or absense^24^. RF and gradient boosting algorithms are less likely to overfit training data than a single decision tree ^25^.

Ten- fold cross-validation was used to train and measure model performance. For CART and RF, there was a 90/10 train-test split in each fold. For XGB and CB, there was a 72/18/10 train-test-validation split in each fold, as 10% was used for validation, and of the resulting 90%, 80% was used for training and the rest for testing. Hyperparameters of XGB and CB were tuned using randomised grid-search prior to model training.

Classification algorithms were trained twice, once with the training data being upsampled and balanced through the use of Synthetic Minority Oversample Technique (SMOTE), and once without. Imbalanced classes, i.e. one class being substantially less represented in a training dataset than others, can lead to poorer classifier accuracy for minority classes ^26^. SMOTE creates synthetic data for the minority class by randomly placing samples along an imaginary line between similar minority samples, with the intention that increasing the number of training instances of a particular class will increase the classifiers sensitivity to them ^27^. In the whole cohort, there were 115 patients that did develop *de novo* HLA-specific antibody post-transplant and 345 that did not. Therefore, SMOTE was applied to the class that did develop *de novo* HLA-specific antibody post-transplant, using synthetic data to increase the size of the class to 345, the same size as the class that did not.

Precision is the proportion of instances of a class which are identified correctly, and recall is the proportion of actual class samples which are identified correctly. F1 score is the harmonic mean of precision and recall. F1 score is a preferred evaluation metric when classes are imbalanced and was therefore calculated to evaluate model performance ^28^. A two-way ANOVA was performed to analyse the effect of classification algorithm and whether SMOTE was used on model F1 score, with Tukey HSD. Of the models trained using the whole cohort, the top three best performing models exhibiting the highest F1 score, were chosen for further evaluation by XAI.

SHapley Additive exPlanation (SHAP) values use a game theoretic approach to calculate the importance of each features contribution to a final model prediction output. For each feature of each training data point, a SHAP value was calculated, where the SHAP value corresponds to how the data point influenced the model output ^29^. A SHAP value of 0 has no impact on model outcome, a negative value impacted the model to predict the outcome as the patient not developing *de novo* HLA-specific antibody, whereas a positive value impacted the model to predict the outcome as the patient developing *de novo* HLA-specific antibody, while the magnitude of the value reflects the strength of this effect. SHAP summary plots and dependence plots were visually inspected to understand how the model attributed each feature’s importance as the value of the feature changed.

Machine learning analysis was performed in Python (version 3.6). The sklearn package was used for CART and RF, and xgboost package for XGB. (catboost 1.0.4; sklearn 0.24.2; xgboost 1.5.1).

Data in figures and text is presented as mean ± standard deviation (SD) unless otherwise specified.

## Results

Four hundred and sixty patients were identified for inclusion in the analysis. Demographic, pre-transplant and post-transplant variables are shown in Table 2.

**Table 2:**
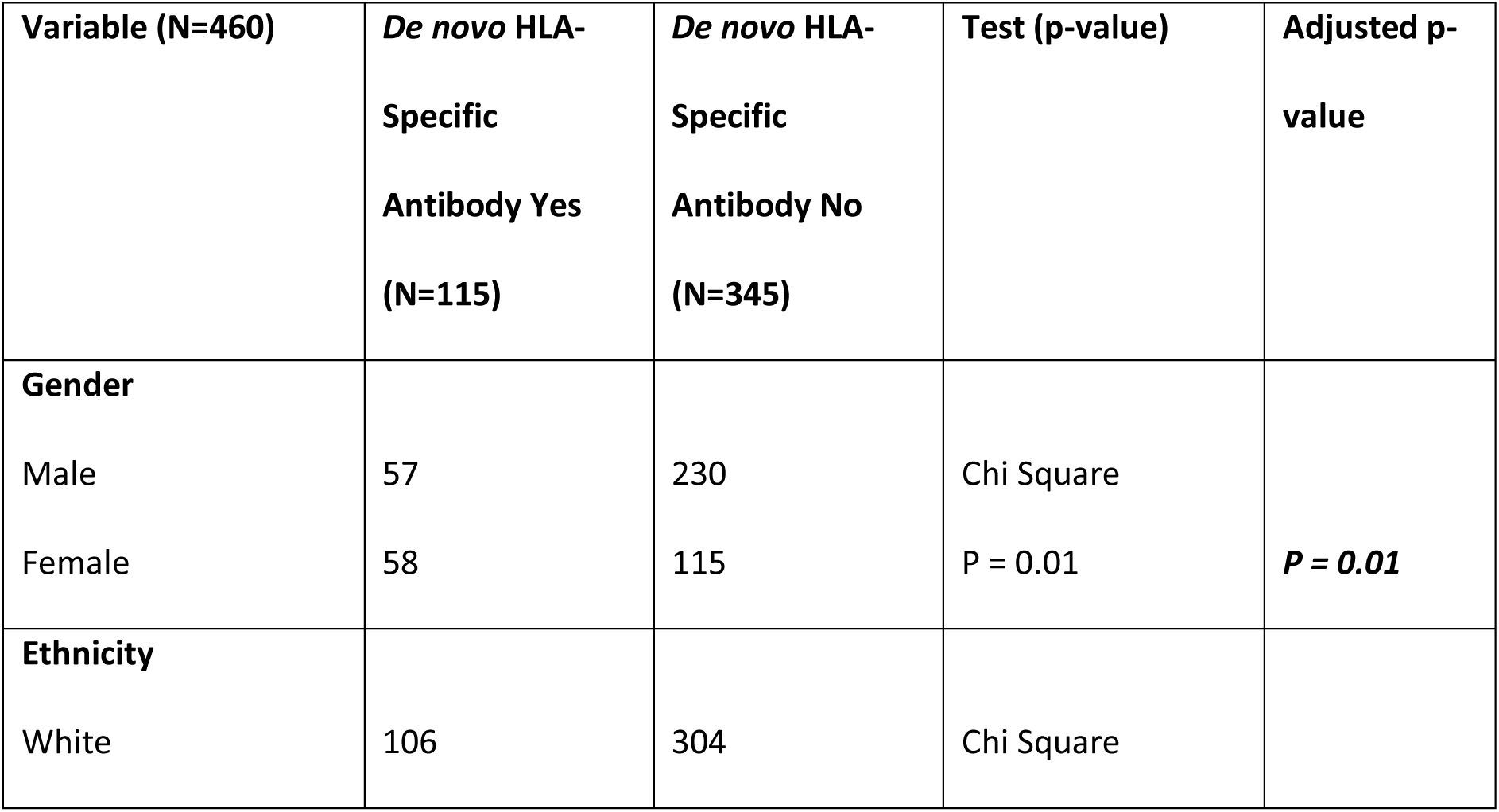
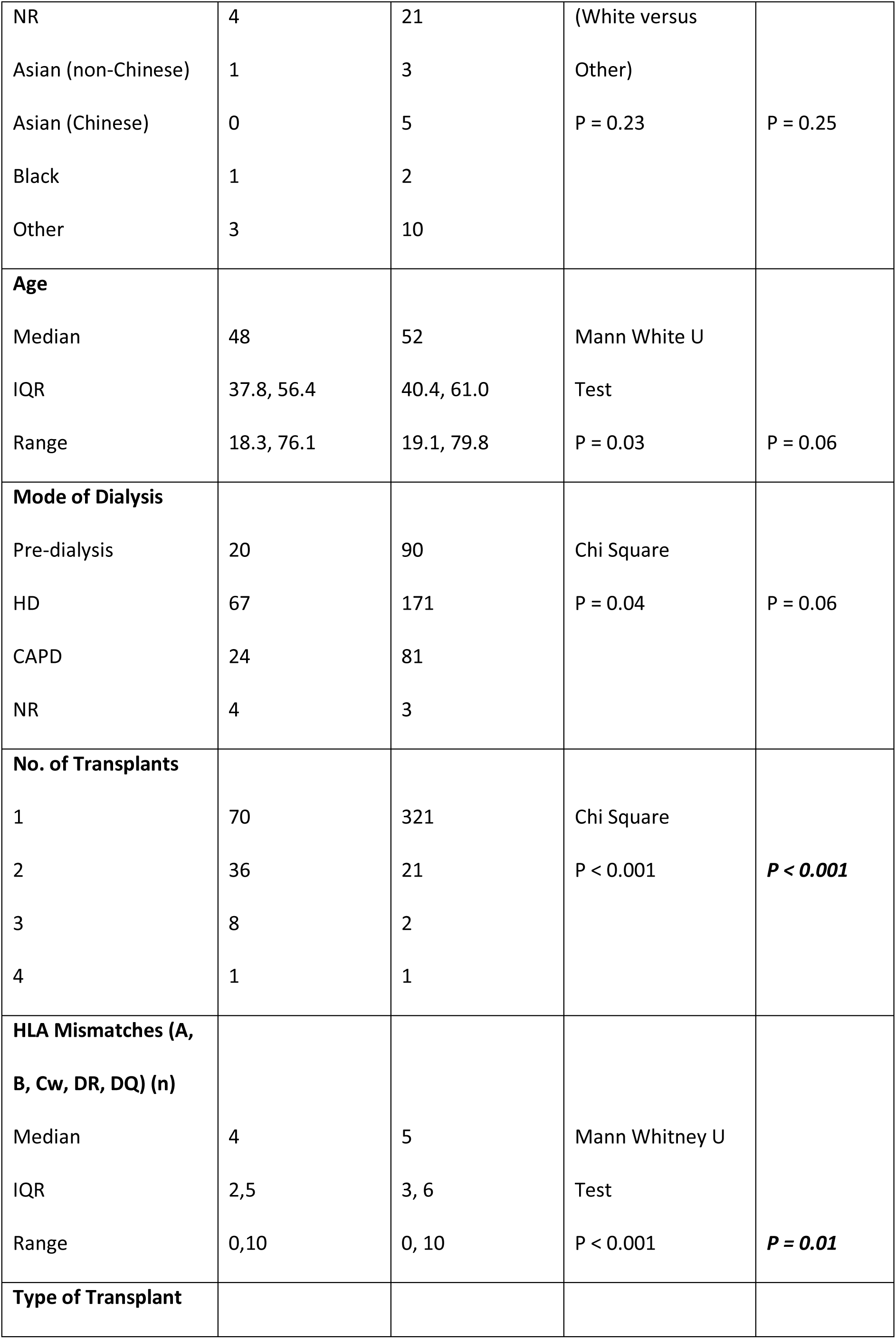
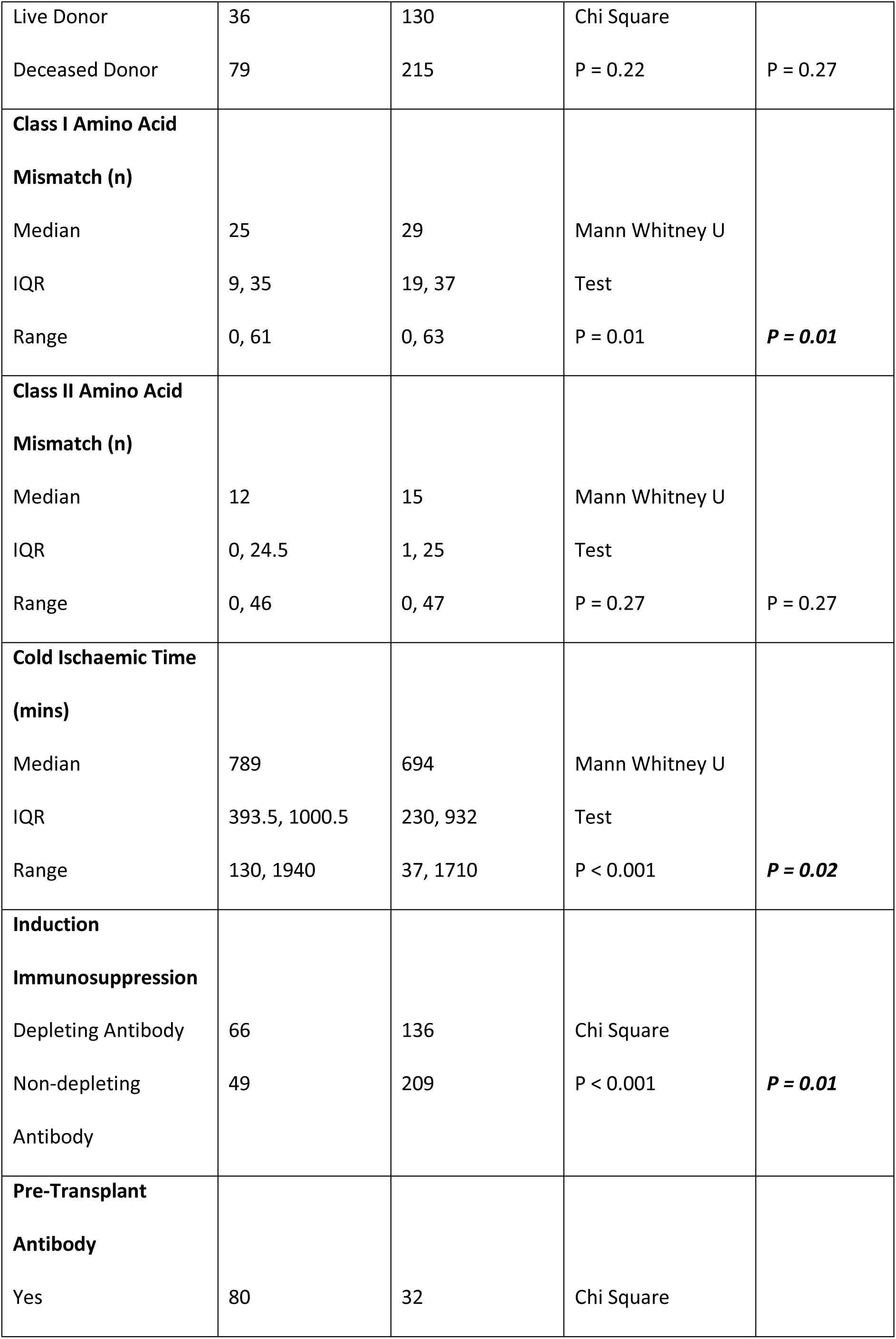
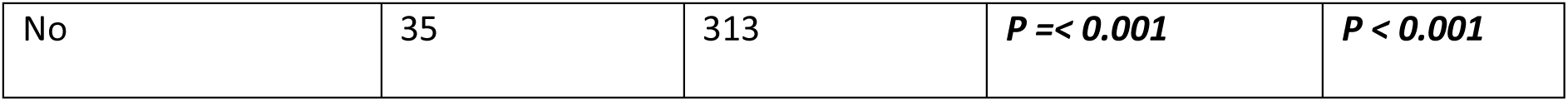
Univariate Analysis for Pre-transplant Variables.

### Subgroup Analysis

As referred to in the methods, a subset analysis was then performed of the same analysis for the cohort of patients who did not have any pre-transplant sensitisation which provided 348 patients for analysis. Univariate analysis is shown in Table 3.

**Table 3:** Subgroup univariate analysis for unsensitised pre-transplant patients for Pre-transplant Variables.

| **Variable (N=348)** | ***De novo* HLA-Specific Antibody Yes (N=35)** | ***De novo* HLA-Specific Antibody No (N=313)** | **Test (p-value)** | **Adjusted p-value** |
| --- | --- | --- | --- | --- |
| **Gender** |  |  |  |  |
| Male | 22 | 215 | Chi Square |  |
| Female | 13 | 98 | P = 0.48 | P = 0.60 |
| **Ethnicity** |  |  |  |  |
| White | 33 | 276 | Chi Square |  |
| NR | 1 | 18 | (White versus |  |
| Asian (non-Chinese) | 0 | 2 | Other) |  |
| Asian (Chinese) | 1 | 5 | P = 0.28 | P = 0.51 |
| Black | 0 | 2 |  |  |
| Other | 0 | 10 |  |  |
| **Age** |  |  |  |  |
| Median | 48 | 52 | Mann White U |  |
| IQR | 35.3, 54.7 | 41.0, 61.2 | Test |  |
| Range | 18.3, 74 | 19.1, 79.8 | *P* < **0.05** | P = 0.30 |
| **Mode of Dialysis** |  |  |  |  |
| Pre-dialysis | 9 | 82 | Chi Square |  |
| HD | 12 | 151 | P = 0.24 | P = 0.51 |
| CAPD | 13 | 77 |  |  |
| NR | 1 | 3 |  |  |
| **No. of Transplants** |  |  |  |  |
| 1 | 30 | 295 | Chi Square |  |
| >1 | 5 | 18 | P = 0.05 | P = 0.30 |
| **HLA Mismatches (A, B, Cw, DR, DQ) (n)** |  |  |  |  |
| Median | 5 | 5 | Mann Whitney U |  |
| IQR | 3, 5.5 | 3, 6 | Test |  |
| Range | 0, 10 | 0, 10 | P = 0.75 | P = 0.82 |
| **Type of Transplant** |  |  |  |  |
| Live Donor | 11 | 117 | Chi Square |  |
| Deceased Donor | 24 | 196 | P = 0.49 | P = 0.60 |
| **Class I Amino Acid Mismatch (n)** |  |  |  |  |
| Median | 30 | 29 | Mann Whitney U |  |
| IQR | 19.5, 37 | 20, 38 | Test |  |
| Range | 0, 53 | 0, 63 | P = 0.91 | P = 0.91 |
| **Class II Amino Acid Mismatch (n)** |  |  |  |  |
| Median | 23 | 17 |  |  |
| IQR | 4, 27.5 | 2, 25 | Mann Whitney U |  |
| Range | 0, 46 | 0, 47 | Test | P = 0.51 |
|  |  |  | P = 0.15 |  |
| **Cold Ischaemic Time**<br>**(mins)** |  |  |  |  |
| Median | 659 | 676 | Mann Whitney U |  |
| IQR | 305, 809.5 | 230, 922 | Test |  |
| Range | 130, 1145 | 37, 1710 | P = 0.39 | P = 0.60 |
| **Induction**<br>**Immunosuppression** |  |  |  |  |
| Depleting Antibody | 17 | 122 | Chi Square |  |
| Non-depleting<br>Antibody | 18 | 191 | P = 0.27 | P = 0.51 |

### Machine Learning

#### Total Patient Cohort Analysis

On models trained and evaluated using data from the entire patient cohort, a two-way ANOVA showed that the classification algorithm had a statistically significant effect on model F1 score (*p* < 0.001), as did using SMOTE to upsample the minority class (*p* = 0.05) (Figure 1). However, the interaction between these terms was not significant (*F*(3, 72) = 0.18, *p* = 0.9).

**Figure 1:**
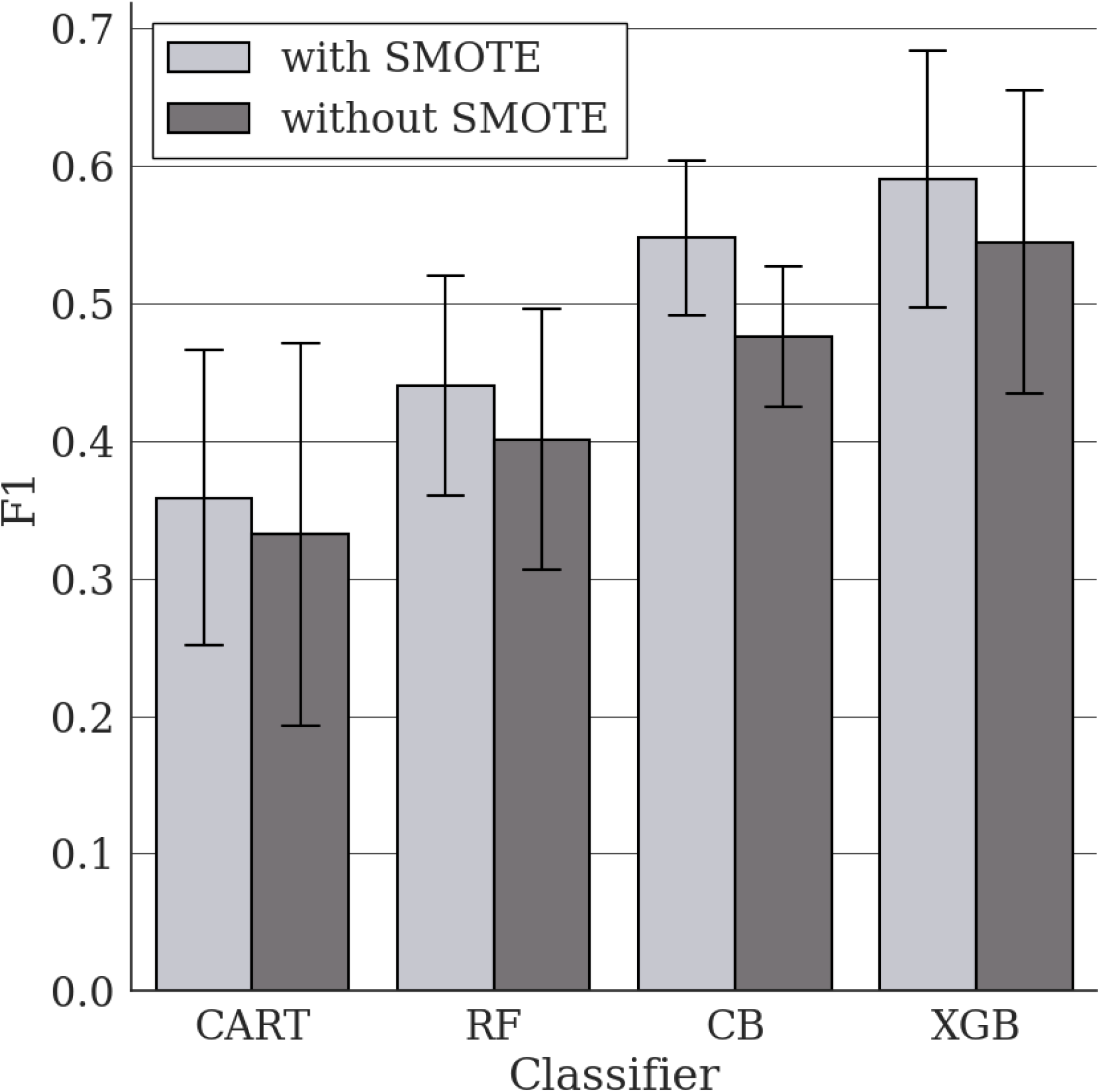
Mean (± SD) F1 score for classifier models on the whole cohort. Models were cross-validated over ten folds containing data from 460 patients. This was repeated twice, once using SMOTE during training to upsample the minority class (those who did develop *de novo* HLA-specific antibody) to the same size as the majority class, and once without.

Tukey post-hoc test revealed significant pairwise differences between the F1 score of models generated by CART and RF algorithms compared to the F1 score of models generated by CB and XGB (CART *vs.* CB: −0.17 F1, CART *vs.* XGB: −0.22 F1, RF *vs.* CB: −0.09 F1, RF *vs*. XGB: −0.15). XGB models were the most accurate (with SMOTE: 0.59 ± 0.09, without SMOTE: 0.54 ± 0.12), followed by CB (with SMOTE: 0.55 ± 0.06, without SMOTE: 0.48 ± 0.05), before RF (with SMOTE: 0.44 ± 0.08, without SMOTE: 0.40 ± 0.09), and CART (with SMOTE: 0.36 ± 0.11, without SMOTE: 0.33 ± 0.15).

Of the models trained and evaluated using entire patient cohort data, the models with the three highest F1 scores were further interrogated. Each of these models was an XGB model, one was trained using SMOTE on the minority class (F1: 0.79), the other two were trained without (F1: 0.74 and 0.72). Feature importance was calculated for each of these models using gain, the average improvement in model accuracy when a feature is included (Figure 2). The two most important features ranked by average increase in gain across the three models were number of pre-transplant HLA-specific antibodies, both Class I and Class II. SHAP dependence plots were visually inspected for the models with the three highest F1 scores to understand how the model attributed each feature’s importance as the value of the feature data changed.

**Figure 2:**
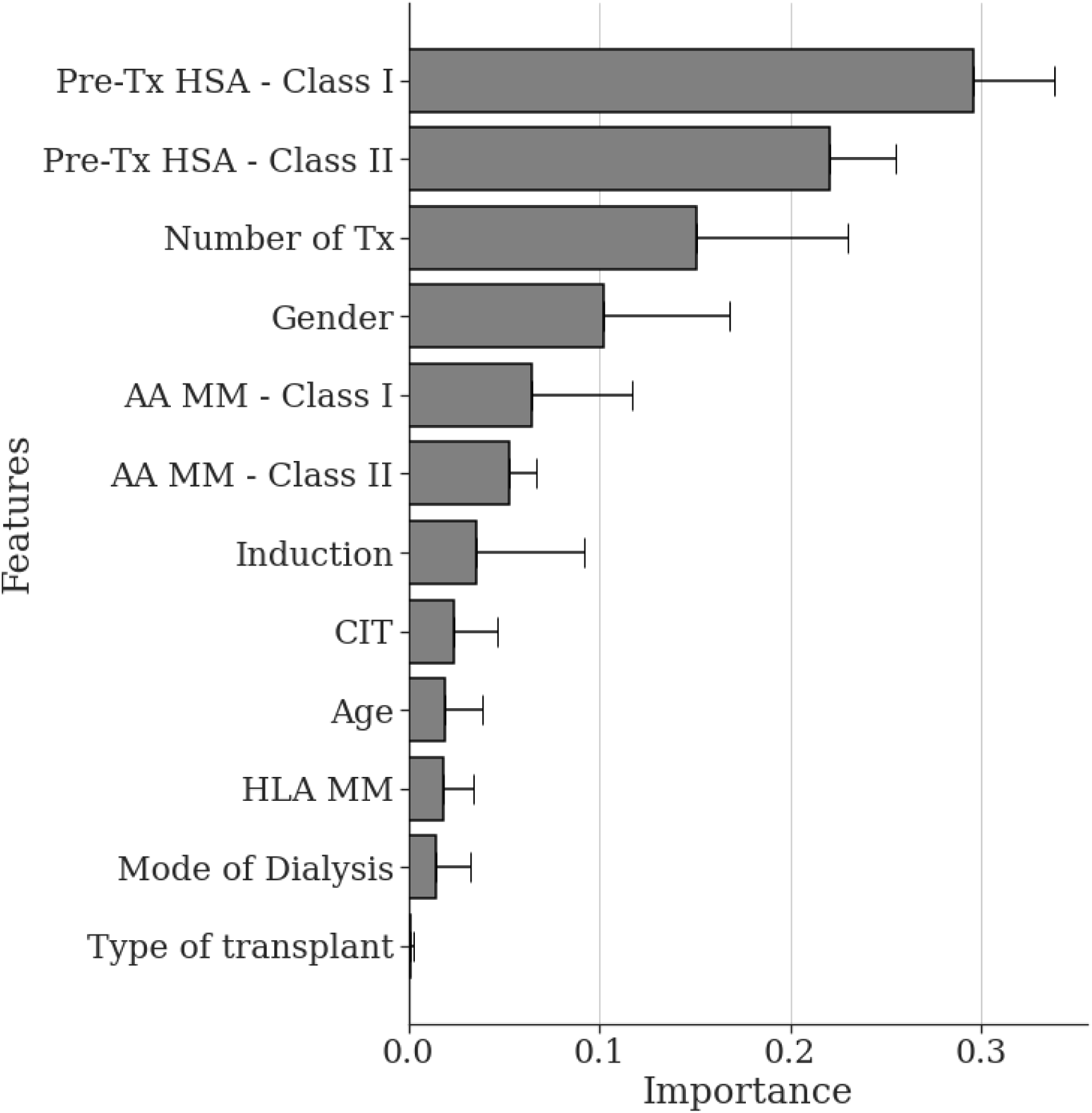
Mean (± SD) gain of each feature across the three most accurate models trained on the whole cohort. Gain is the average improvement in model accuracy when a feature is included in the model and is a measure of the importance of that feature to the model.

#### Pre-transplant HLA Specific Antibody

SHAP dependence plots suggest a relationship between the number of pre-transplant HLA specific antibodies, both Class I and Class II, and the SHAP value of this data (Figure 3). When any pre-transplant HLA-specific antibodies are identified i.e. the count is greater than 0, the associated SHAP values are greater than 0, indicating the model was influenced to predict that the patient did develop post-transplant HLA-specific antibodies. Inversely, when no pre-transplant HLA-specific antibodies were present, i.e. the count was 0, then the associated SHAP values were less than 0, indicating the model was influenced to predict that the patient did not develop post-transplant HLA-specific antibodies.

**Figure 3:**
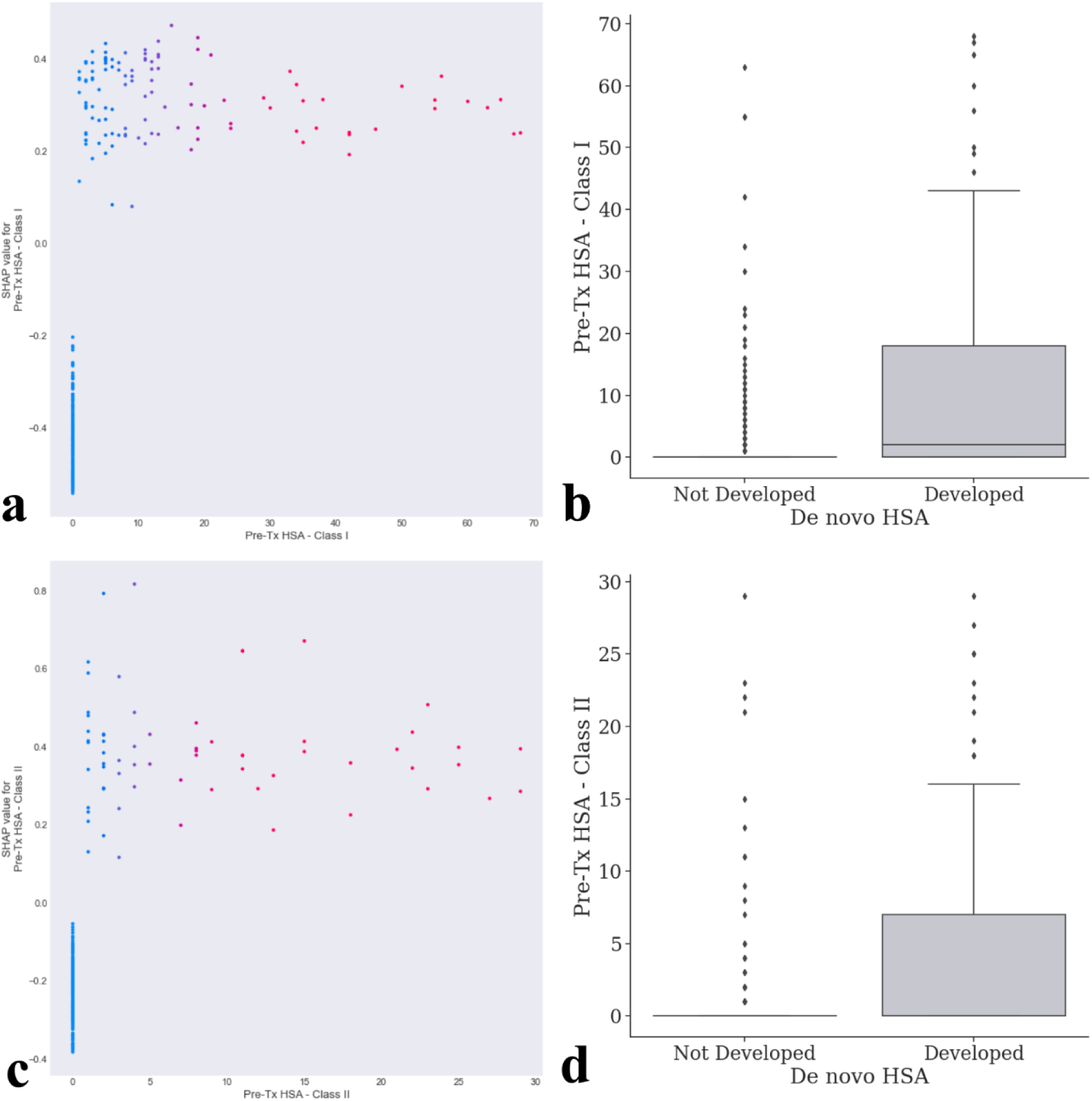
a and c) Example SHAP dependence plots from one of the three best performing models for Pre-transplant HLA Specific Antibody counts (Class I and Class II, respectively) vs SHAP value for Pre-transplant HLA Specific Antibody (Class I and Class II), showing how Pre-transplant HLA Specific Antibody, influences model output. SHAP values of 0, have no impact on model output, SHAP values of <0 indicate the model was influenced to predict that the patient did develop HLA-specific antibody post-transplant, however SHAP values of >0 indicate the model was influenced to predict the inverse. Only training data that was used for that model is plotted, synthetic data generated by SMOTE was not plotted. b and d) boxplots plots showing Pre-transplant HLA Specific Antibody (Class I and Class II) data, each data point and the overall data for the whole cohort, separated by those who did and those who did not develop *de novo* HLA-specific antibody. Horizontal line in the boxplot represents the median, the box limits represent the interquartile range (IQR), whiskers represent 1.5 x IQR, with points outside this range independently plotted.

#### Number of Kidney Transplants

The number of kidney transplants the patient had undergone was the third most important feature in the top three most accurate models when ranked by gain. When the number of kidney transplants the patient had received was 1 i.e. the kidney transplant they received was their only transplant, the SHAP value was less than 0, indicating the model was influenced to predict that these patients did not develop post-transplant *de novo* HLA-specific antibody (Figure 4). Inversely, if they had received a previous transplant, the SHAP values were greater than 0 indicating the model was influenced to predict those patients did develop *de novo* HLA-specific antibody post-transplant.

**Figure 4:**
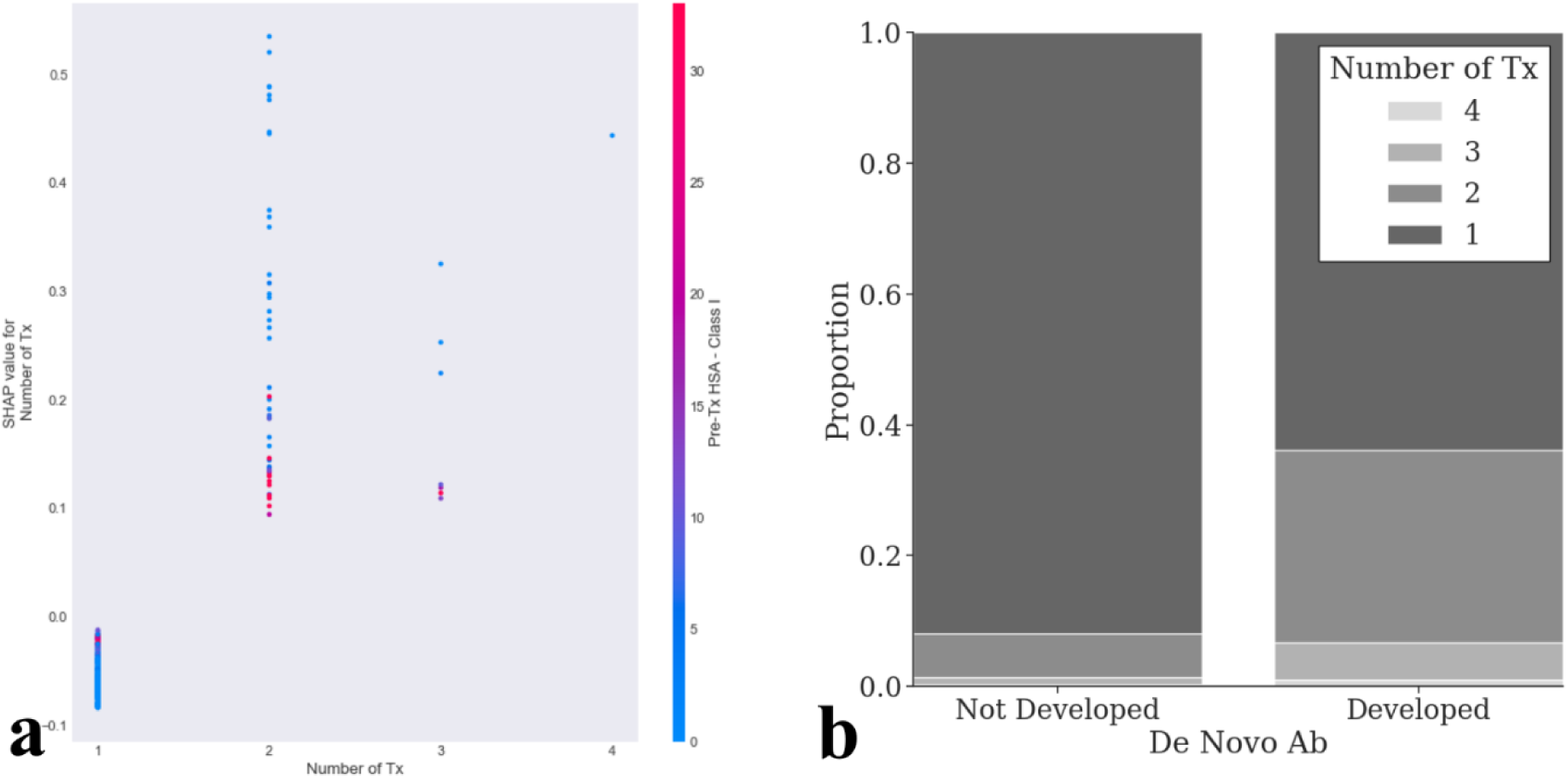
a) An example SHAP dependence plot from one of the three best performing models for Number of Kidney Transplants vs SHAP value for Number of Kidney Transplants, showing how Number of Transplants influences model output. SHAP values of 0, have no impact on model output, SHAP values of <0 indicate the model was influenced to predict that the patient did develop HLA-specific antibody post-transplant, however SHAP values of >0 indicate the model was influenced to predict the inverse. The point of the colour represents the number of Pre-transplant HLA specific antibody – Class I, the feature that approximately had the strongest interaction with Number of Kidney Transplants. Only training data that was used for that model is plotted, synthetic data generated by SMOTE was not plotted. b) boxplots plots showing Number of Kidney Transplants data, each data point and the overall data for the whole cohort, separated by those who did and those who did not develop *de novo* HLA-specific antibody. Horizontal line in the boxplot represents the median (in both cases 0), the box limits represent the interquartile range (IQR), whiskers represent 1.5 x IQR, with points outside this range independently plotted.

#### Cold Ischaemic Times

SHAP dependence plots of CIT showed a similar effect across two of the three best performing models. Figure 5 shows a dependence plot of one of the models where shorter CIT, typically seen in living donors, resulted in negative SHAP values, influencing the model to predict that the patient did not develop HLA-specific antibodies.

**Figure 5:**
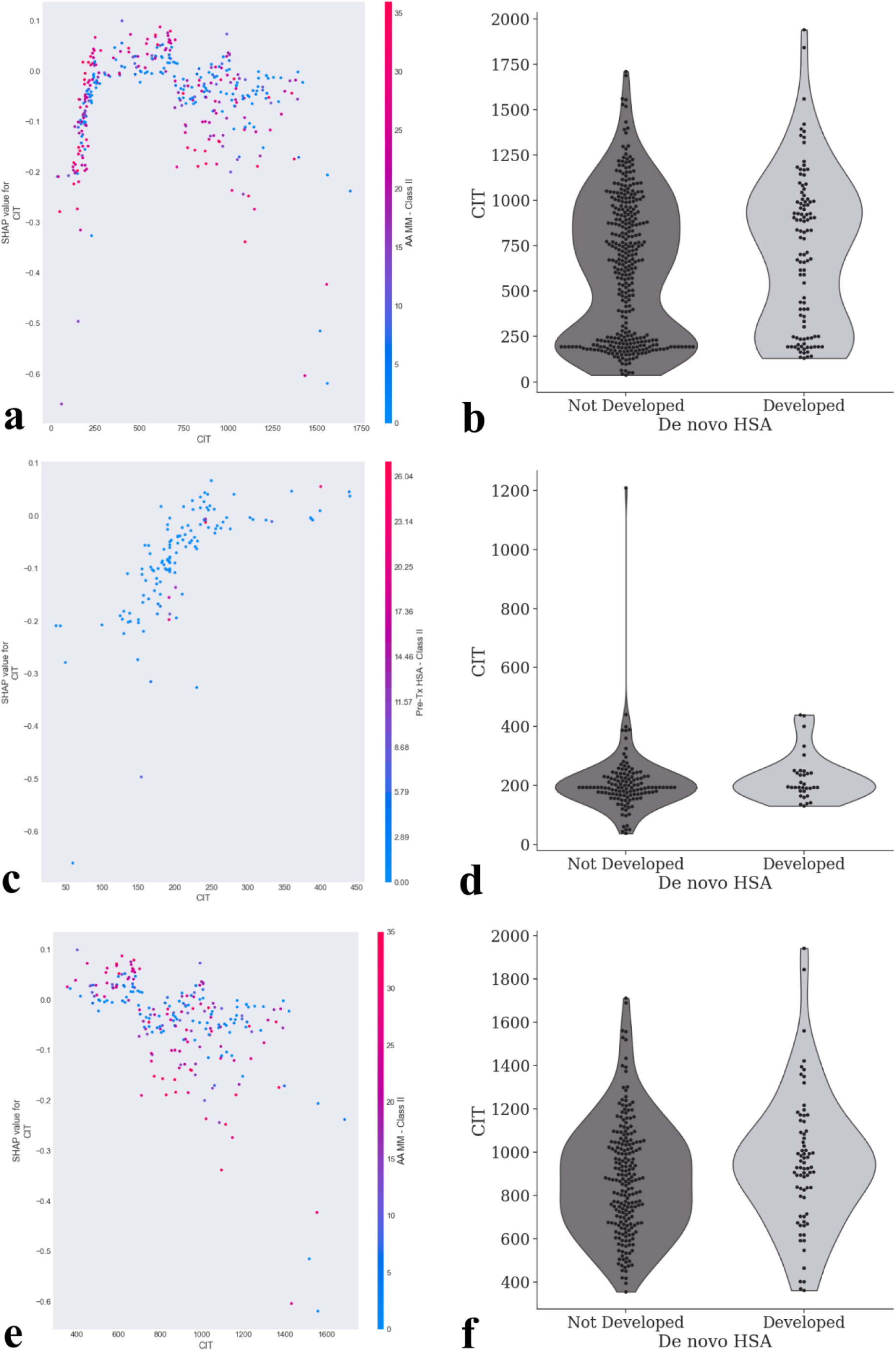
a) An example SHAP dependence plot from one of the three best performing models for Cold Ischaemic Time (mins) vs SHAP value for Cold Ischaemic Time (mins), showing how Cold Ischaemic Time influences model output. SHAP values of 0, have no impact on model output, SHAP values of >0 indicate the model was influenced to predict that the patient did develop HAL-specific antibody post-transplant, however SHAP values of <0 indicate the model was influenced to predict the inverse. The point of the colour represents the number of Class II Amino Acid mismatches, the feature that approximately had the strongest interaction with Cold Ischaemic Time. Only training data that was used for that model is plotted, synthetic data generated by SMOTE was not plotted. b) violin plots showing Cold Ischaemic Time data, each data point and the overall distribution of data for the whole cohort, separated by those who did and those who did not develop *de novo* HLA-specific antibody. c) and d) show the same plots as a) and b), respectively, though showing only data from living donors, whereas e) and f), again show the same plots, however with only data from cadaveric donors.

As CIT increased in the living donors, so did SHAP value. However, longer CIT, typically seen in cadaveric donors, also resulted in negative SHAP values as CIT increased which is somewhat unexpected. Therefore, overall, the shortest and longest CIT times resulted in influencing the models to predict that patients did not develop *de novo* HLA-specific antibody post-transplant.

#### Age

Dependence plots of age showed a similar effect to CIT, where both the extreme ranges of young and old were associated with the models predicting that the individual would not develop HLA-specific antibody post-transplant. The predominant effect, however, was for older adults over the age of 65 (Figure 6) who appear to develop less *de novo* HLA-specific antibody post-transplant.

**Figure 6:**
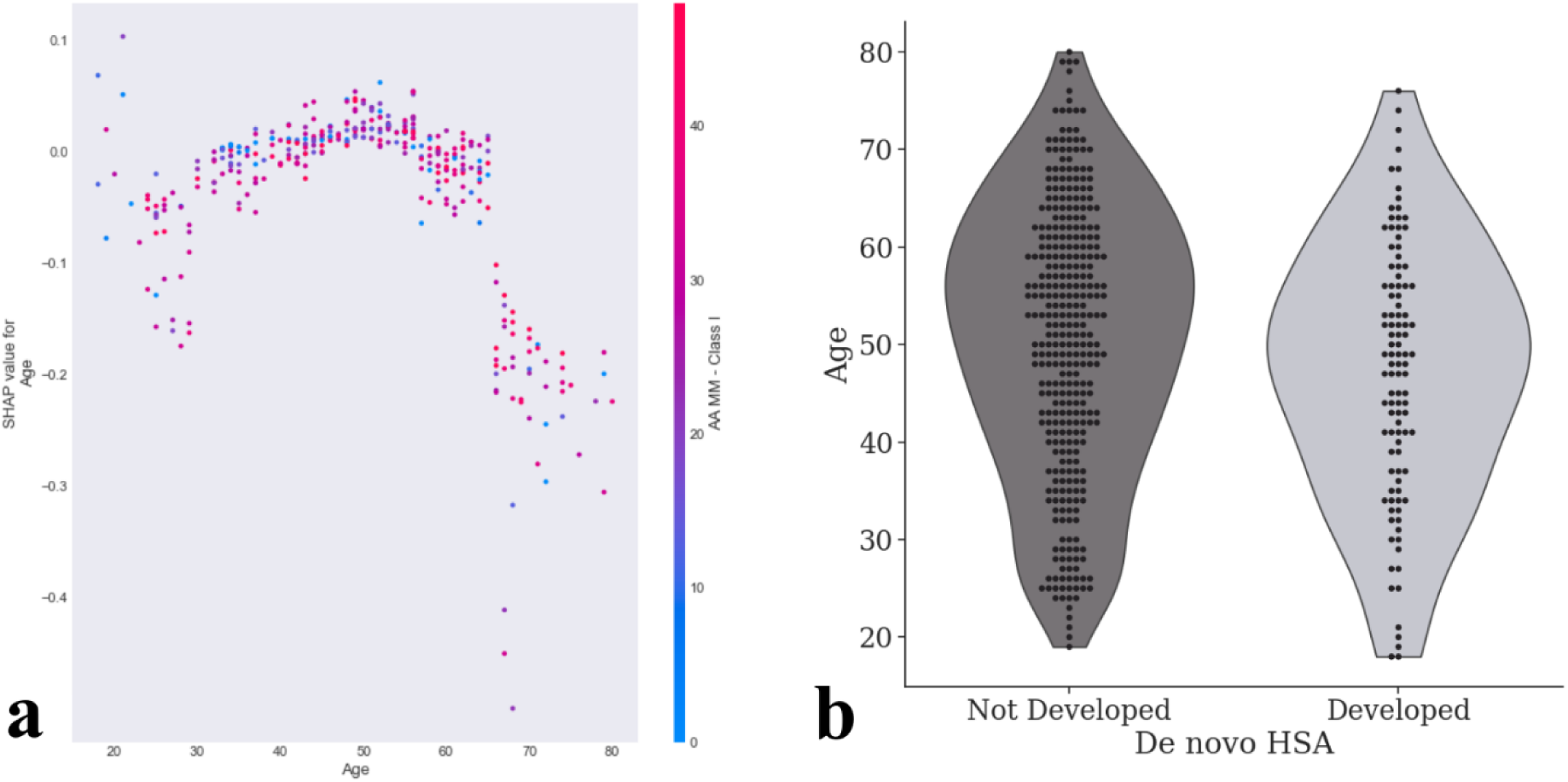
a) An example SHAP dependence plot from one of the three best performing models for Age vs SHAP value for Age, showing how Age influences model output. SHAP values of 0, have no impact on model output, SHAP values of >0 indicate the model was influenced to predict that the patient did develop HLA-specific antibody post-transplant, however SHAP values of <0 indicate the model was influenced to predict the inverse. The point of the colour represents the number of Class I Amino Acid mismatches, the feature that approximately had the strongest interaction with Age. Only training data that was used for that model is plotted, synthetic data generated by SMOTE was not plotted. b) violin plots showing Age data, each data point and the overall distribution of data for the whole cohort, separated by those who did and those who did not develop *de novo* HLA-specific antibody.

#### Gender

Despite females being significantly more likely to develop *de novo* HLA-specific antibody post-transplant (*P=0.01*), SHAP dependence plots showed no clear patterns suggesting gender was influencing model output. Figure 7 shows the proportion of males and females who did and did not develop *de novo* HLA-specific antibody post-transplant.

**Figure 7:**
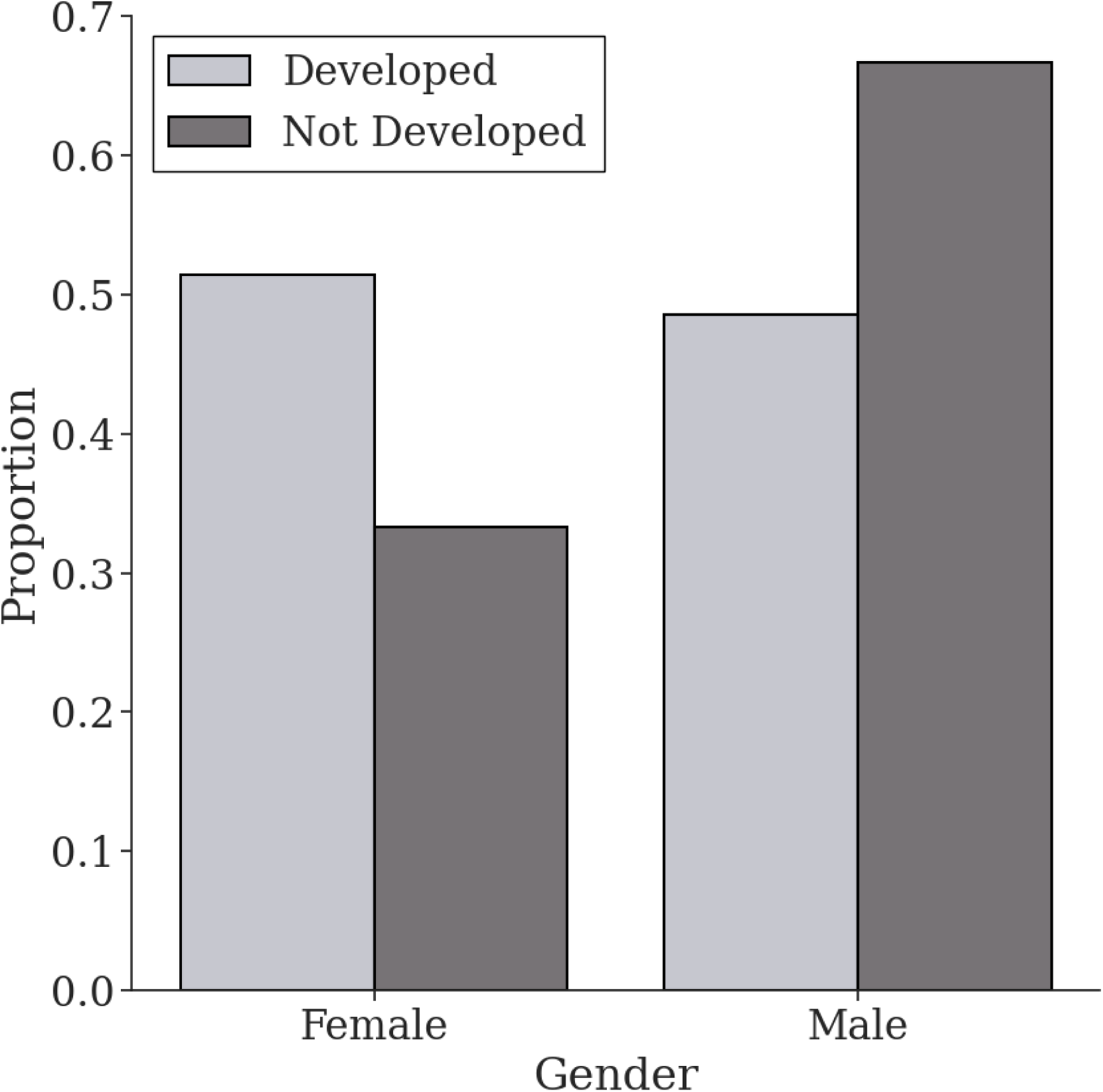
Proportion of patients in the whole cohort (460 patients) who did and did not develop *de novo* HLA-specific antibody, separated by gender.

#### Subgroup Analysis

Models trained using only the unsensitised subgroup had low F1 scores (0.15 ± 0.15 - across all models, with and without the use of SMOTE), therefore these models were not further interrogated using XAI as they could not be trusted to return a reliable prediction (Supplementary Figure S1).

## Discussion

The aim of this study was to examine pre-transplant factors that affect *de novo* HLA-specific antibody development following kidney transplant. This was performed through the use of univariate statistics and interrogating trained machine learning classifier models using XAI. The most important factor positively influencing formation of post-transplant HLA-specific antibody was the presence of pre-transplant HLA-specific antibody and was consistent across both univariate statistics and XAI outcomes. The reasons for this are multifactorial but possible explanations include the semi-quantitative nature of SAB testing and lack of reproducibility of MFIs ^30^; natural and dynamic variation in circulating HLA-specific antibody ^31^ and patients who have had prior sensitising events being primed to develop more HLA-specific antibody. Machine learning models trained in the unsensitised cohort were inaccurate and unreliable, suggesting that the presence of pre-transplant HLA-specific antibody is the most reliable clinical marker.

Also appearing to generate a significant effect on the development of post-transplant HLA-specific antibody was the number of kidney transplants that a patient had received. Patients for whom the kidney transplant was their first transplant were significantly less likely to develop post-transplant HLA-specific antibody. SHAP values reflected this, with any previous transplants pushing the models to predict that the patient would develop *de novo* HLA-specific antibody. This observation is possibly unsurprising since prior transplantation is a significant sensitising event and will correlate closely with the presence of pre-transplant HLA-specific antibody. Similarly, females were significantly more likely to develop *de novo* HLA-specific antibody than males presumably due to their higher exposure to sensitising events in the context of pregnancy, childbirth and miscarriage.

A smaller, but notable, effect was observed with recipient age. Those who were older were less likely to develop post-transplant *de novo* HLA-specific antibody, though this was not significant. This is probably due to immunosenescence which is observed as a dysfunctional and less responsive immune system with advancing age ^32^ and raises the question about the value of immunosuppression minimisation in this cohort to mitigate against side effects and infection ^33^.

CIT was significantly lower in the group that did not develop post-transplant HLA-specific antibody. It is a complex, bimodal variable which is illustrated in the SHAP plot (Figure 5) and from which we can make some observations. Firstly, we observe longer CIT for living donors was associated with the model predicting post-transplant *de novo* HLA-specific antibody development. This is probably due to longer live donor CITs being observed with transplants in the kidney sharing scheme which are normally retrieved at a site distant to the implanting hospital, and indicate a degree of immunological complexity by nature of being in the scheme. Another factor increasing CIT in the context of living kidney donation is surgical complexity which could indicate a second or subsequent transplant and its association with pre-transplant HLA-specific antibody. Next, we observed that longer CIT in cadaveric donation was associated with the model predicting that post-transplant *de novo* HLA-specific antibody would not be developed. This observation is less obvious to rationalise but is most prominent at the longest CIT which may indicate a late offer that would only be accepted for an excellent graft and a straightforward recipient from the surgical and immunological perspective aiming to achieve the best possible graft outcomes.

Patients who received a non-depleting induction immunosuppressant were significantly less likely to develop post-transplant *de novo* HLA-specific antibody. In our centre, non-depleting induction therapy is only used for standard immunological risk transplants, so any high-risk transplant including re-transplants and those with pre-transplant HLA-specific antibody would receive depleting induction therapy. Thus, a depleting induction agent can be considered to be a surrogate for pre-transplant sensitisation.

Several groups have suggested that where there is serological evidence of non-DSA *de novo* HLA-specific antibody, that DSA may be bound to the transplanted kidney thereby reducing the serum load, making it undetectable on serological testing ^34–37^. In our view, an important next step would be to perform a paired serum and intra-graft biopsy study (DSA elution and histological features of rejection) to examine whether non-DSA *de novo* HLA-specific antibody is, indeed, a marker for intra-graft DSA, with or without biopsy and clinical evidence of rejection.

This paper demonstrates how the combined use of ML and SHAP presents a powerful framework for firstly, modelling complex relationships, and secondly, identifying the key factors influencing these relationships. In the present study, the identification of the complex relationships CIT and age have with post-transplant *de novo* HLA-specific antibody formation was only possible due to the use of these methods. While caution should be exercised in interpreting these results, due to the limitations of SHAP, further discussed below, this paper demonstrates how this method enhances data interpretation beyond that possible with basic statistical tests.

### Limitations

In our study, the number of patients developing HLA-specific antibody were a reasonable proportion of the total cohort, but interpretation of the unsensitised subgroup was limited due to the small numbers of *de novo* HLA-specific antibody formed in these patients. Pre-transplant HLA-specific antibody appears to be the predominant predictor of post-transplant HLA-specific antibody, but with the caveats described of assay cut-off, MFI variability and lack of reproducibility introducing bias and raising the question of what is actually *de novo*.

While univariate statistics generally supported the findings and patterns identified by SHAP dependence plots, it is worth noting that SHAP interpretations are only as useful as the accuracy of the models. To mitigate this, only the best performing models were interrogated using XAI. Secondly, analysis took place using only data derived from a single-centre and using relatively few patients. Future research should explore these relationships using a larger cohort.

## Conclusion

In conclusion, we have demonstrated that pre-transplant sensitisation is the biggest predictor of post-transplant *de novo* HLA-specific antibody formation. We have reasonable evidence that female gender, induction immunosuppression and a history of previous transplants are other factors which influence post-transplant antibody formation, though our study cannot draw robust conclusions regarding DSA formation due to the small sample size with notable effects seen in age and CIT. In an unsensitised cohort, machine learning models are imprecise, but multivariate analysis indicates age, transplant type and cold ischaemic time being predominant pre-transplant factors influencing post-transplant *de novo* HLA-specific antibody formation.

## Data Availability Statement

Code for running experiments is available on GitHub at (https://github.com/CBFLivUni/ML_Kidney_Tx). Data may be shared upon reasonable request to the corresponding author.

## Acknowledgements

This work was supported by Kidney Research Northwest (formerly Mersey Kidney First) Grant Number 42/13

