## Supplement for "Using Machine Learning to Examine Pre-Transplant Factors Influencing *De novo* HLA-Specific Antibody Development Post-Kidney Transplant"

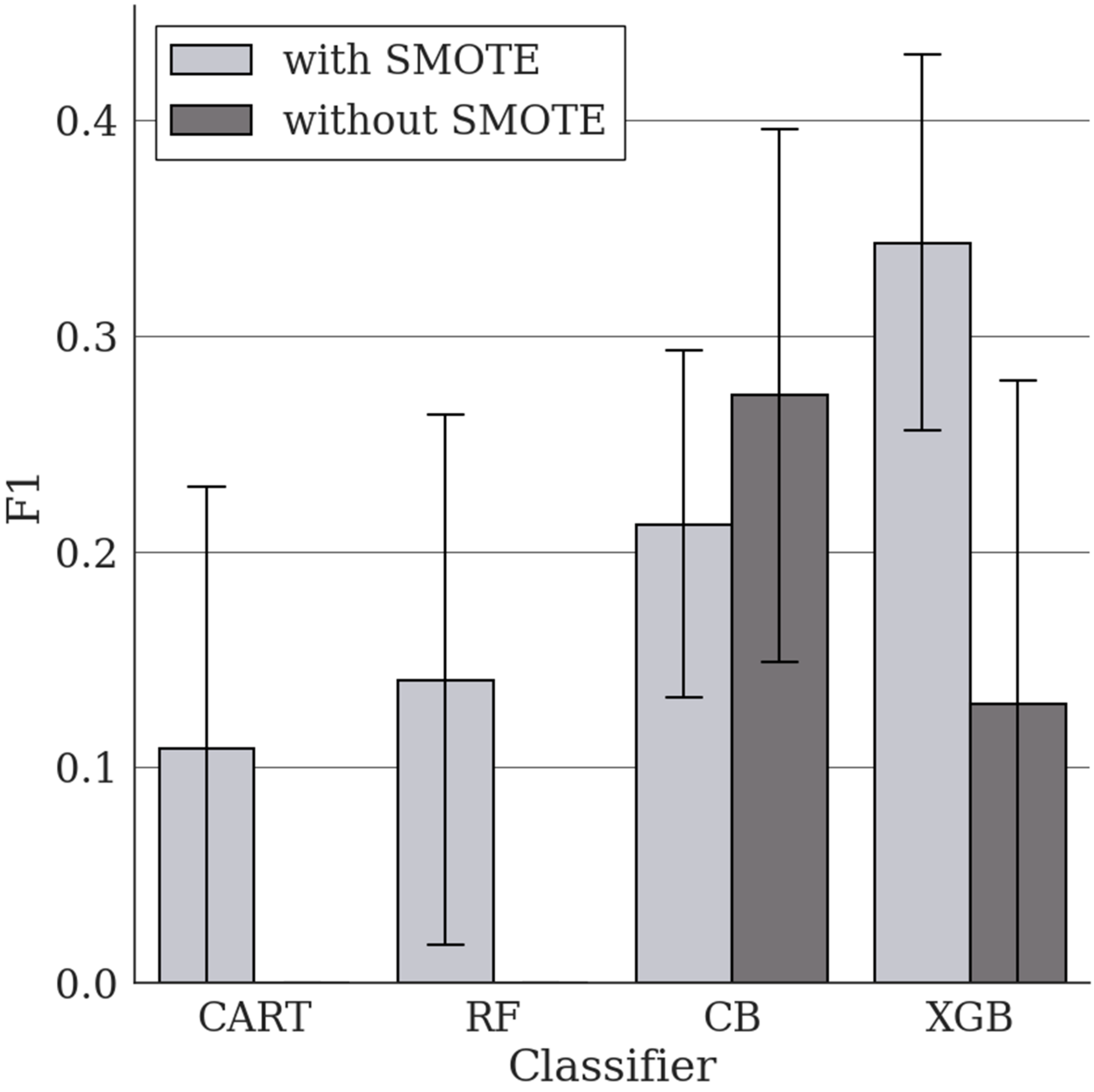


**Supplementary Figure S1** Mean (± SD) F1 score for classifier models on the unsensitised cohort. Models were cross-validated over ten folds containing data from 348 patients. This was repeated twice, once using SMOTE during training to upsample the minority class (those who did develop *de novo* HLA-specific antibody) to the same size as the majority class, and once without. Mean and SD for CART and RF models without SMOTE was 0.

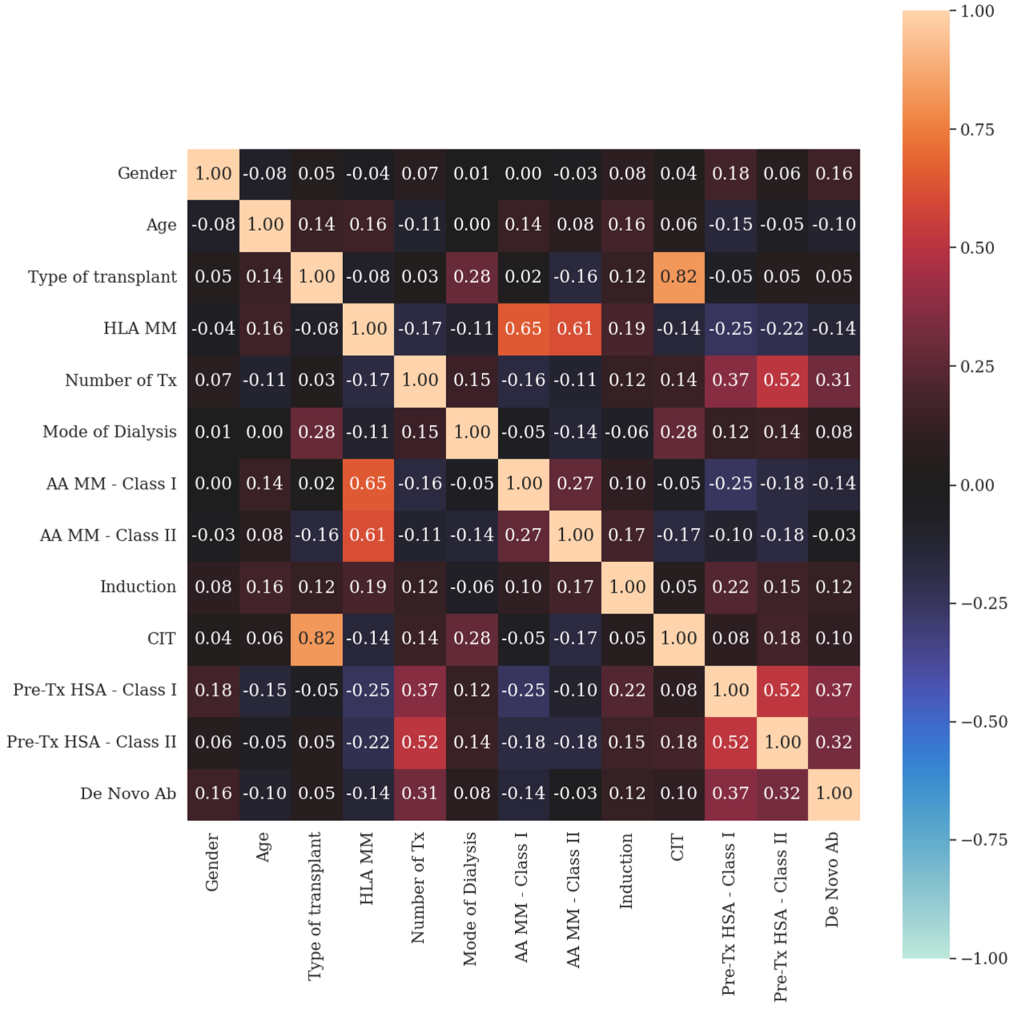


**Supplementary Figure S2.** Correlation between all pre-transplant variables and *de novo* HLA-specific antibody development post-transplant. The number and shading represents Pearson's R for continuous-to-continuous variable comparisons, correlation ratio for categorical-to-continuous variable comparisons and Cramer's V for categorical-to-categorical variable comparisons.
